# Determining population-level allocation strategies for COVID-19 treatments in the United States using a quantitative framework, a case study using nirmatrelvir/ritonavir

**DOI:** 10.1101/2022.08.04.22278431

**Authors:** Alexandra Savinkina, Gregg Gonsalves, Joseph S. Ross, A. David Paltiel

## Abstract

**Background:** New COVID-19 medications force decision makers to weigh limited evidence of efficacy and cost in determining which patient populations to target for treatment. A case in point is nirmatrelvir/ritonavir, a drug that has been recommended for elderly, high-risk individuals, regardless of vaccination status, even though clinical trials have only evaluated it in unvaccinated patients. A simple optimization framework might inform a more reasoned approach to the tradeoffs implicit in the treatment allocation decision.

**Methods:** We used a mathematical model to analyze the cost-effectiveness of four nirmatrelvir/ritonavir allocation strategies, stratified by vaccination status and risk for severe disease. We considered treatment effectiveness at preventing hospitalization ranging from 21% to 89%. Sensitivity analyses were performed on major parameters of interest. A web-based tool was developed to permit decision-makers to tailor the analysis to their settings and priorities.

**Results:** Providing nirmatrelvir/ritonavir to unvaccinated patients at high-risk for severe disease was cost-saving when effectiveness against hospitalization exceeded 33% and cost-effective under all other data scenarios we considered. The cost-effectiveness of other allocation strategies, including those for vaccinated adults and those at lower-risk for severe disease, depended on willingness-to-pay thresholds, treatment cost and effectiveness, and the likelihood of severe disease.

**Conclusions:** Priority for nirmatrelvir/ritonavir treatment should be given to unvaccinated persons at high-risk of severe disease from COVID-19. Further priority may be assigned by weighing treatment effectiveness, disease severity, drug cost, and willingness to pay for deaths averted.

## Introduction

More than two years into the COVID-19 pandemic, the U.S. is still experiencing hundreds of COVID-19 deaths a day [1]. In January 2022, COVID-19 was among the top four leading causes of death in the US for every age group, and was the top cause of death for those over age 45 [2]. Fortunately, vaccines and other medical treatments have reduced the severity of COVID-19 infection in both hospitalized and non-hospitalized patients. Through March 21, 2022, vaccines alone have averted an estimated 2.3 million deaths in the U.S., saving the country nearly $900 billion [3].

Alongside vaccination, several effective treatments for COVID-19 have been developed. One of the more promising is Pfizer’s 5-day oral antiviral treatment regimen of nirmatrelvir/ritonavir (brand name Paxlovid), granted emergency use authorization by the U.S. Food and Drug Association (FDA) in December 2021. In clinical trials, nirmatrelvir/ritonavir showed an 89% reduction in hospitalizations and no deaths in high-risk unvaccinated COVID-19 positive patients [4]. Such a reduction in disease severity could ease strain on scarce hospital and critical care resources. Other treatments have shown more modest effects in this setting and more treatments continue to be developed [5].

Given both the speed with which new therapeutic agents are being developed and the continuing urgency of the COVID-19 pandemic, decision makers will inevitably and repeatedly be to asked to make approval and coverage decisions, long before the clinical and economic impacts of these treatment options are fully understood. On the basis of its current price and observed efficacy, the FDA has given emergency use authorization to nirmatrelvir/ritonavir for treatment of the elderly and other high-risk adults, regardless of vaccination status [6]. Some observers have questioned the breadth of this decision, noting that other studies have failed to replicate the effectiveness observed in the initial trial [7–10] and that nirmatrelvir/ritonavir’s efficacy remains unproven in vaccinated patients [11, 12], in addition to the risks posed by serious drug interaction and toxicity [13] and the uncertainty of rebound infections and symptoms [14, 15]. Still others have wondered whether the substantial reduction in risks of hospitalization and death in patients receiving nirmatrelvir/ritonavir might justify expanding the indications for treatment far beyond the highest-risk patient population [16].

We sought to provide practical guidance to clinicians, policy-makers, and payers regarding the clinical, epidemiological, and economic circumstances under which a new medication of uncertain efficacy might serve as a cost-effective and appropriate use of COVID-19 treatment resources across a range of different target populations. The significance of our analysis lies less in the particular application to nirmatrelvir/ritonavir and more in the elaboration of a generalized evaluation framework and the establishment of performance benchmarks for future COVID-19 treatment options, many of which are likely to display therapeutic properties against SARS-CoV-2 infection.

## Methods

### Model structure

We used a decision tree model to analyze the effectiveness and cost-effectiveness of different allocation strategies of nirmatrelvir/ritonavir in the United States (Figure 1). The target population of the model includes those who are newly COVID-19 positive (COVID-19+) within the timeframe eligible for nirmatrelvir/ritonavir prescription (within 5 days of positive test or onset of symptoms) [4]. Individuals are assigned a probability of being at high-risk vs low-risk for severe COVID-19, a probability of being vaccinated for COVID-19, a probability of hospitalization dependent on risk and vaccination status, and a probability of death if hospitalized.

**Figure 1.**
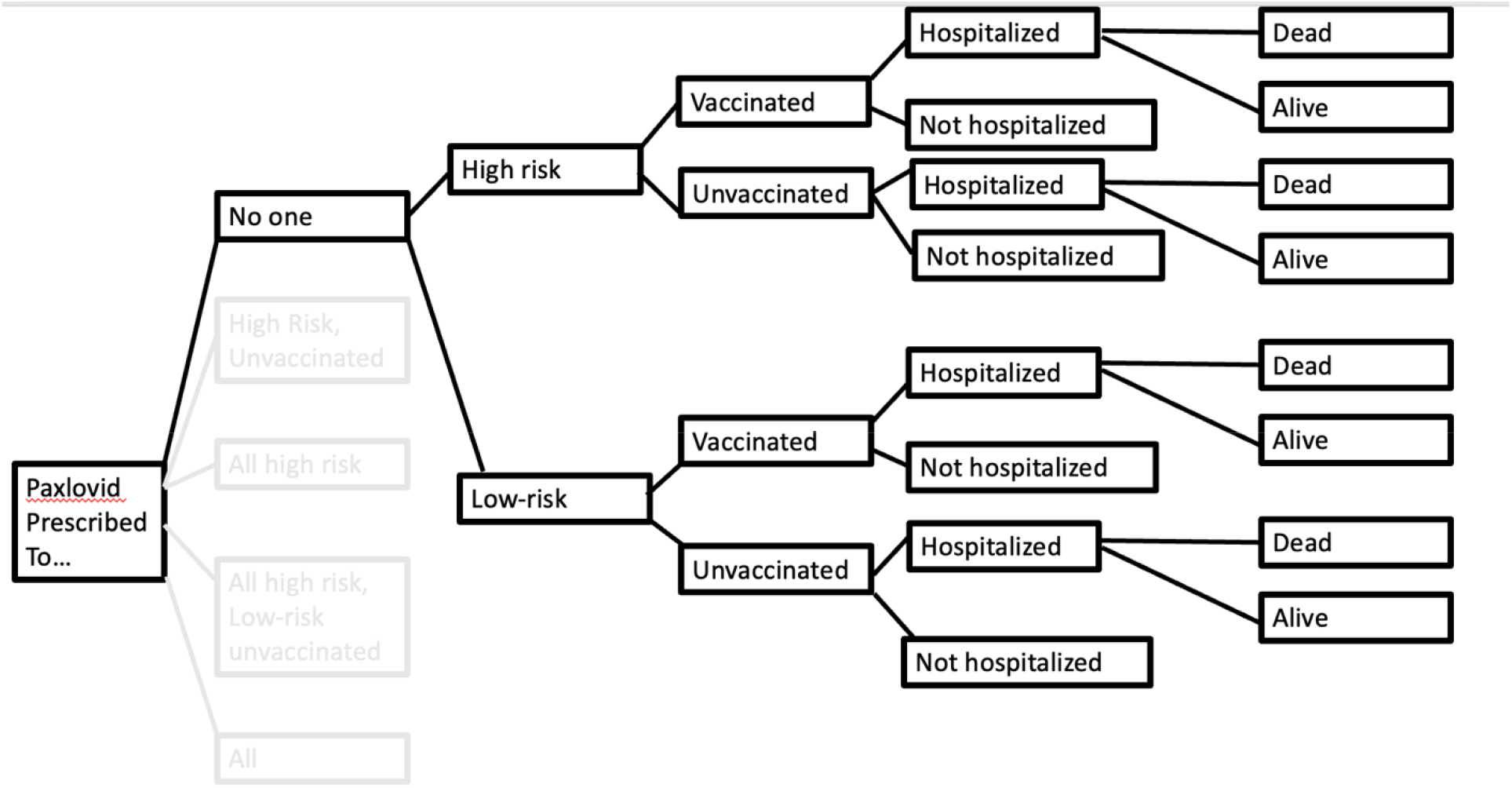
Model structure.

### Strategies

In addition to the baseline policy of treating nobody with nirmatrelvir/ritonavir, we considered 4 increasingly expansive eligibility policies for persons with confirmed SARS-CoV-2 infection:

1. Unvaccinated patients at high-risk for severe COVID-19;
2. All patients at high-risk for severe COVID-19, regardless of vaccination status;
3. All unvaccinated patients and vaccinated patients at high-risk for severe COVID-19; and
4. All patients

### Model parameter values

“High-risk” for severe COVID-19 was determined according to inclusion criteria for Pfizer’s nirmatrelvir/ritonavir trial [4], which included everyone over 60 years of age or with at least one risk factor for severe COVID-19 [6]. Vaccination rates came from US data on vaccination rates nationwide [1]. Only primary series vaccinations were considered for both vaccination rates and vaccination effect. We present analyses using a range of nirmatrelvir/ritonavir treatment effect modifiers on hospitalization: 89% from Pfizer’s nirmatrelvir/ritonavir trial [4], and 21%-67% from more recent literature [7–10].

The only costs considered in this model were the cost of a course of nirmatrelvir/ritonavir in the US [17], and the cost of a COVID-19 hospitalization in the United States, which was estimated using published literature [18, 19]. We did not consider other costs as they were not expected to differ between scenarios, and therefore were not expected to alter our results or conclusions.

All model parameter values and ranges used in sensitivity analysis can be found in Table 1.

**Table 1.**
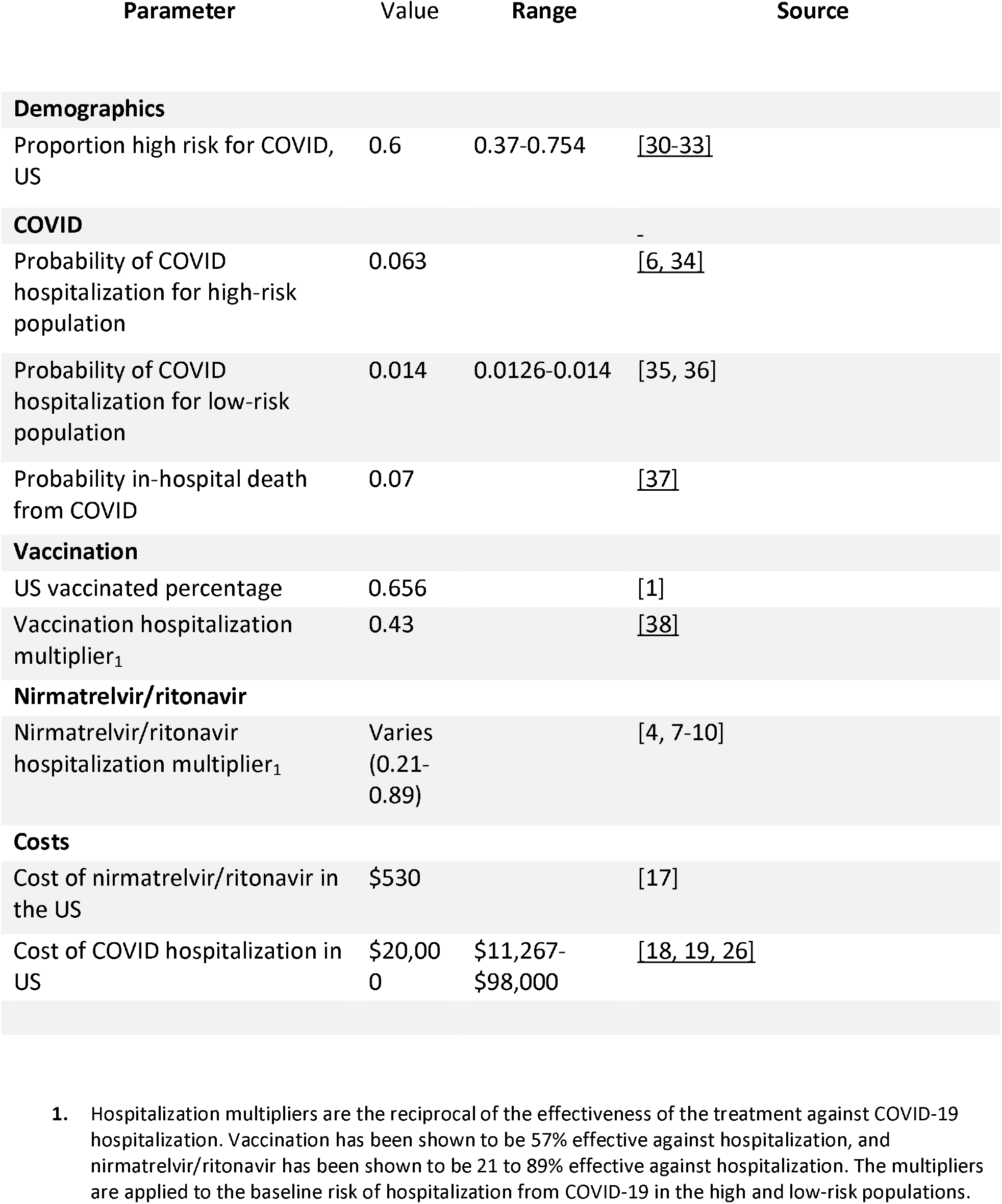
Model parameter values and ranges.

| Parameter | Value | Range | Source |
| --- | --- | --- | --- |
| **Demographics** |  |  |  |
| Proportion high risk for COVID, US | 0.6 | 0.37-0.754 | [30-33] |
| **COVID** |  |  |  |
| Probability of COVID hospitalization for high-risk population | 0.063 |  | [6, 34] |
| Probability of COVID hospitalization for low-risk population | 0.014 | 0.0126-0.014 | [35, 36] |
| Probability in-hospital death from COVID | 0.07 |  | [37] |
| **Vaccination** |  |  |  |
| US vaccinated percentage | 0.656 |  | [1] |
| Vaccination hospitalization multiplier<sub>1</sub> | 0.43 |  | [38] |
| **Nirmatrelvir/ritonavir** |  |  |  |
| Nirmatrelvir/ritonavir hospitalization multiplier<sub>1</sub> | Varies (0.21-0.89) |  | [4, 7-10] |
| **Costs** |  |  |  |
| Cost of nirmatrelvir/ritonavir in the US | \$530 | | [17] |
| Cost of COVID hospitalization in US | \$20,000 | \$11,267-\$98,000 | [18, 19, 26] |

1. Hospitalization multipliers are the reciprocal of the effectiveness of the treatment against COVID-19 hospitalization. Vaccination has been shown to be 57% effective against hospitalization, and nirmatrelvir/ritonavir has been shown to be 21 to 89% effective against hospitalization. The multipliers are applied to the baseline risk of hospitalization from COVID-19 in the high and low-risk populations.

### Economic performance measures

We modeled cost-effectiveness from a healthcare sector perspective, considering the cost of a hospitalization for COVID-19 in the US as well as the cost of nirmatrelvir/ritonavir itself.

Effectiveness measures considered were decreases in risks of hospitalization and death. Incremental cost-effectiveness ratios (ICERs) were measured in dollars per hospitalization averted and per death averted [20].

Net monetary benefit [21] of each strategy was also considered under a variety of willingness-to-pay thresholds, ranging from $10,000 per death averted to $5 million per death averted. Net monetary benefit was calculated by multiplying the incremental benefit of each strategy as compared to no nirmatrelvir/ritonavir by the willingness to pay per death averted, and then subtracting out the cost of the strategy.

We considered value of a statistical life (VSL) estimates as a way to think about willingness to pay threshholds per death averted [23].

### Sensitivity analyses

We conducted several one-way sensitivity analyses, varying key parameters to their highest or lowest range, as reported in Table 1. Parameters analyzed included average cost of US COVID-19 hospitalization, nirmatrelvir/ritonavir effectiveness in vaccinated individuals and low-risk individuals, and risk of hospitalization from COVID-19.

In addition, we conducted a two-way sensitivity analysis, identifying the preferred allocation as a function of both nirmatrelvir/ritonavir treatment effectiveness (0 to 100% effectiveness at preventing hospitalization) and cost of nirmatrelvir/ritonavir treatment course (0 to $2,000, allowing for the need for a potential second course as well as variation in treatment price).

We also developed a <u>publicly available tool</u> which can be used to vary any model parameter and determine the best allocation strategy when assessed using the net monetary benefit approach. This tool can be used to reproduce all analyses reported here. It can also be used to evaluate alternative treatment strategies with differing effectiveness measures, as well as treatment strategies in different populations, in a range of alternative clinical and economical circumstances.

## Results

All results are reported on a per-eligible-person basis. In the status quo scenario with no treatment (Strategy 0), average population risk of hospitalization was 0.0268, risk of death was 0.00187, and cost was $536 (Supplemental Table 1A, Supplemental Table 2A).

Offering nirmatrelvir/ritonavir to high-risk unvaccinated COVID+ patients (Strategy 1) was cost-saving when nirmatrelvir/ritonavir effectiveness against hospitalization was assumed to exceed 33% (Table 2, Table 3). At 33% effectiveness against hospitalization, the cost per hospitalization averted was $5,000 and the cost per death averted was $70,900. When nirmatrelvir/ritonavir effectiveness against hospitalization was assumed to be 21%, the ICER for hospitalization averted was $19,500 and for death averted was $279,000.

**Table 2.** Incremental cost-effectiveness ratios for hospitalizations prevented by nirmatrelvir/ritonavir. By differing allocation scenarios, and presented for different effectiveness at preventing hospitalization estimates for nirmatrelvir/ritonavir from the literature.

|  | Incremental cost-effectiveness ratios (ICER) per hospitalization averted, according to given nirmatrelvir/ritonavir effectiveness at preventing hospitalization measure |  |  |  |  |
| --- | --- | --- | --- | --- | --- |
|  | 89% effective, Hammond et al.[4] | 67% effective, Arbel et al.[10] | 45% effective, Dryden-Peterson et al.[9] | 33% effective, Mefsin et al.[7] | 21% effective, Yip et al.[8] |
| **Strategy 0: No nirmatrelvir/ritonavir** |  |  |  |  |  |
| **Strategy 1 - nirmatrelvir/ritonavir for unvaccinated high-risk** | Cost saving | Cost-Saving | Cost-Saving | \$5,000 | \$19,500 |
| **Strategy 2 - nirmatrelvir/ritonavir for all high-risk (regardless of vaccination)** | \$700 | \$7,600 | \$21,400 | \$36,600 | \$69,300 |
| **Strategy 3 - nirmatrelvir/ritonavir all high risk and unvaccinated low-risk** | \$26,700 | \$42,300 | \$72,900 | \$106,900 | \$179,800 |
| **Strategy 4 - nirmatrelvir/ritonavir for all** | \$88,600 | \$124,400 | \$195,300 | \$273,800 | \$441,900 |

**Table 3.** Incremental cost-effectiveness ratios for deaths prevented by nirmatrelvir/ritonavir. By differing allocation scenarios, and presented for different effectiveness at preventing hospitalization estimates for nirmatrelvir/ritonavir from the literature.

|  | Incremental cost-effectiveness ratios (ICER) per death averted, according to given nirmatrelvir/ritonavir effectiveness at preventing hospitalization measure |  |  |  |  |
| --- | --- | --- | --- | --- | --- |
|  | 89% effective, Hammond et al.[4] | 67% effective, Arbel et al.[10] | 45% effective, Dryden-Peterson et al.[9] | 33% effective, Mefsin et al.[7] | 21% effective, Yip et al.[8] |
| **Strategy 0: No nirmatrelvir/ritonavir** |  |  |  |  |  |
| **Strategy 1 - nirmatrelvir/ritonavir for unvaccinated high-risk** | Cost saving | Cost-Saving | Cost-Saving | \$70,900 | \$279,000 |
| **Strategy 2 - nirmatrelvir/ritonavir for all high-risk (regardless of vaccination)** | \$9,500 | \$108,900 | \$305,500 | \$523,200 | \$989,800 |
| **Strategy 3 - nirmatrelvir/ritonavir all high risk and unvaccinated low-risk** | \$381,900 | \$603,600 | \$1,042,100 | \$1,527,600 | \$2,568,200 |
| **Strategy 4 - nirmatrelvir/ritonavir for all** | \$1,265,600 | \$1,777,500 | \$2,789,900 | \$3,911,000 | \$6,313,500 |

Offering nirmatrelvir/ritonavir to high-risk COVID+ patients regardless of vaccination (Strategy 2) led to costs per hospitalization averted ranging from $700 to $69,300 and costs per death averted ranging from $9,500 to $989,800 for effectiveness assumptions ranging from 89% to 21%.

Using the same range of effectiveness assumptions (89% to 21%), we found that offering nirmatrelvir/ritonavir to all high-risk patients and to unvaccinated low-risk patients (Strategy 3) would have costs per hospitalization averted ranging from $26,700 to $179,800 and costs per deaths averted of $381,900 to $2,568,200. Treating all patients (Strategy 4) yielded costs per hospitalization averted ranging from $88,600 to $441,900 and costs per deaths averted of $1,265,600 $6,313,500.

Some studies have shown significantly reduced effectiveness of nirmatrelvir/ritonavir in low-risk patients [10]. When we reduced the effectiveness of nirmatrelvir/ritonavir against hospitalization in these patients to one third of that seen in high-risk patients, costs per hospitalization averted increased threefold to fivefold for Strategies 3 and 4 (Supplemental Table 3).

Clinical trials of nirmatrelvir/ritonavir effectiveness have yet to be conducted in vaccinated patients. When we assumed a 50% reduction in nirmatrelvir/ritonavir effectiveness in vaccinated patients, costs per hospitalization doubled in Strategy 2 and more than tripled in Strategy 4 (Supplemental Table 4).

When we lowered our assumption of the average US cost of a COVID-19 hospitalization from $20,000 to $10,000, the cost per death averted increased in every strategy, though providing nirmatrelvir/ritonavir to high-risk vaccinated people remained cost-saving when the assumed nirmatrelvir/ritonavir effectiveness against hospitalization was 89% (Supplemental Table 5). While dropping the cost of hospitalization lowered the cost of a COVID-19 infection in every scenario, it also raised the percentage of that cost attributable to nirmatrelvir/ritonavir treatment.

To explore treatment cost-effectiveness under a potentially more severe viral variant, we doubled the risk of hospitalization. Costs per hospitalization averted were roughly halved in every strategy (Supplemental Table 6). Providing nirmatrelvir/ritonavir to both high-risk unvaccinated and high-risk vaccinated people became cost-saving with a high nirmatrelvir/ritonavir effectiveness against hospitalization.

Figures 2 and 3 report the optimal allocation strategy, under different willingness to pay threshholds, as both nirmatrelvir/ritonavir cost and effectiveness were varied across their plausible ranges.

**Figure 2.**
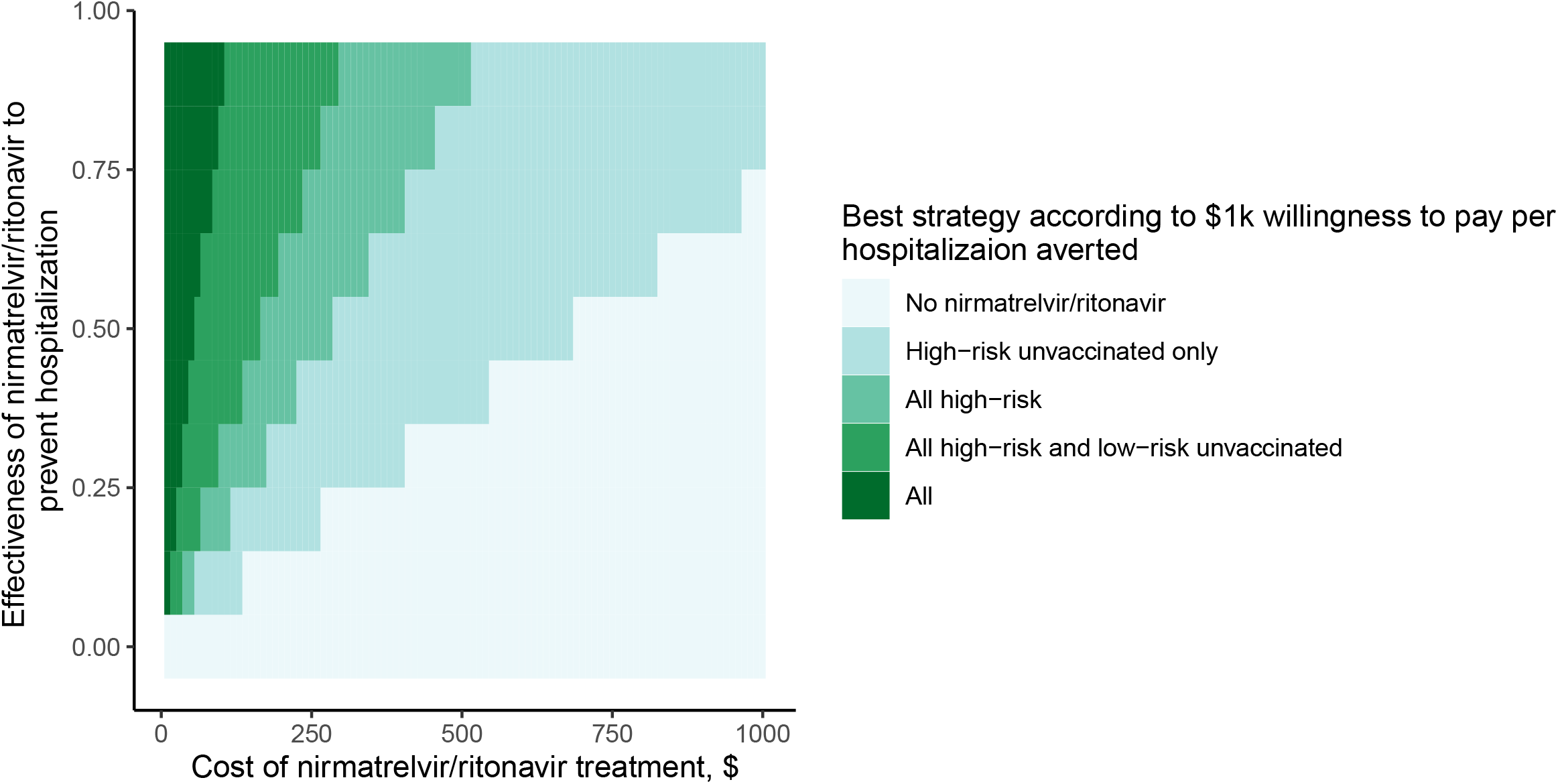
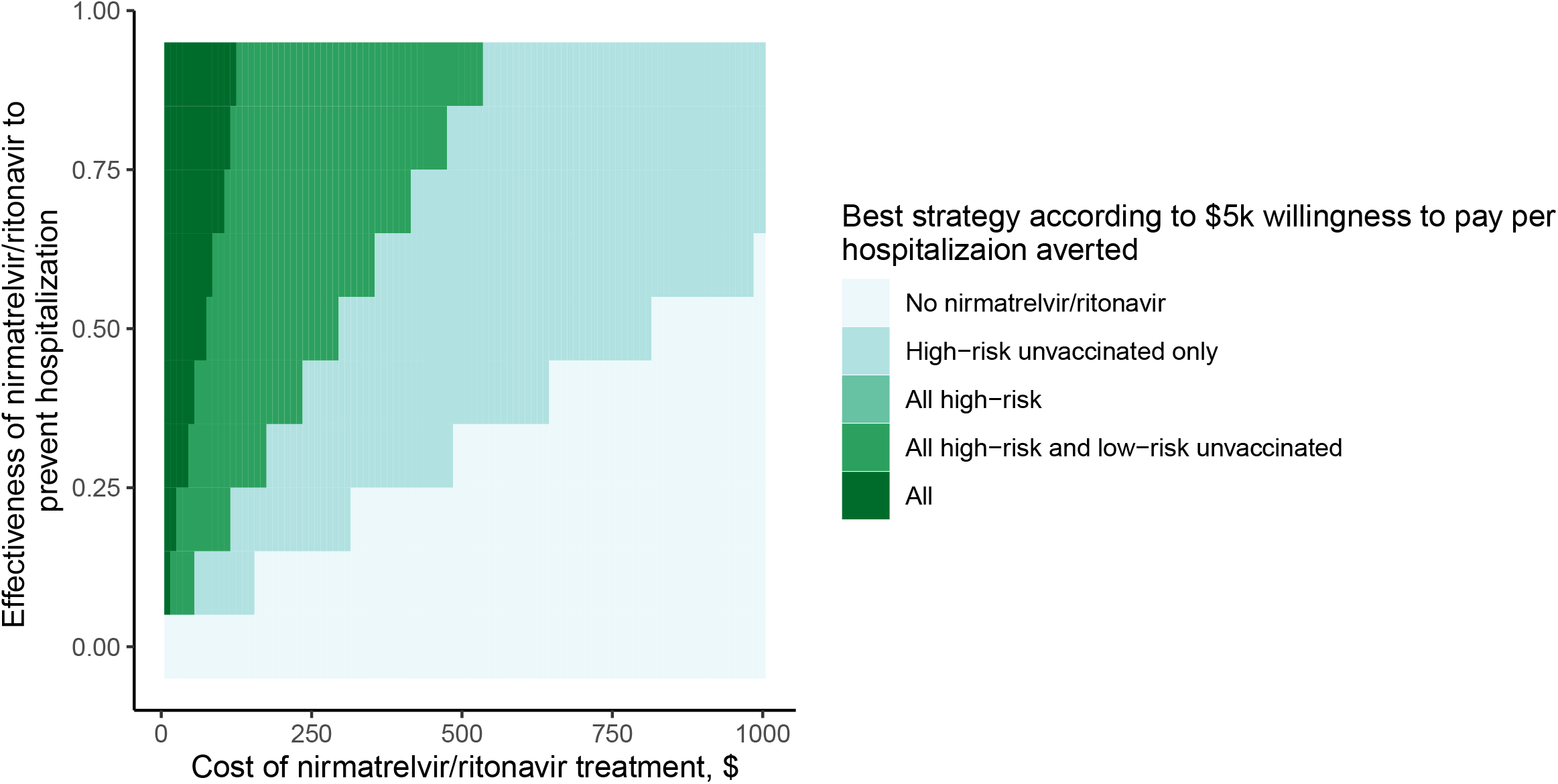
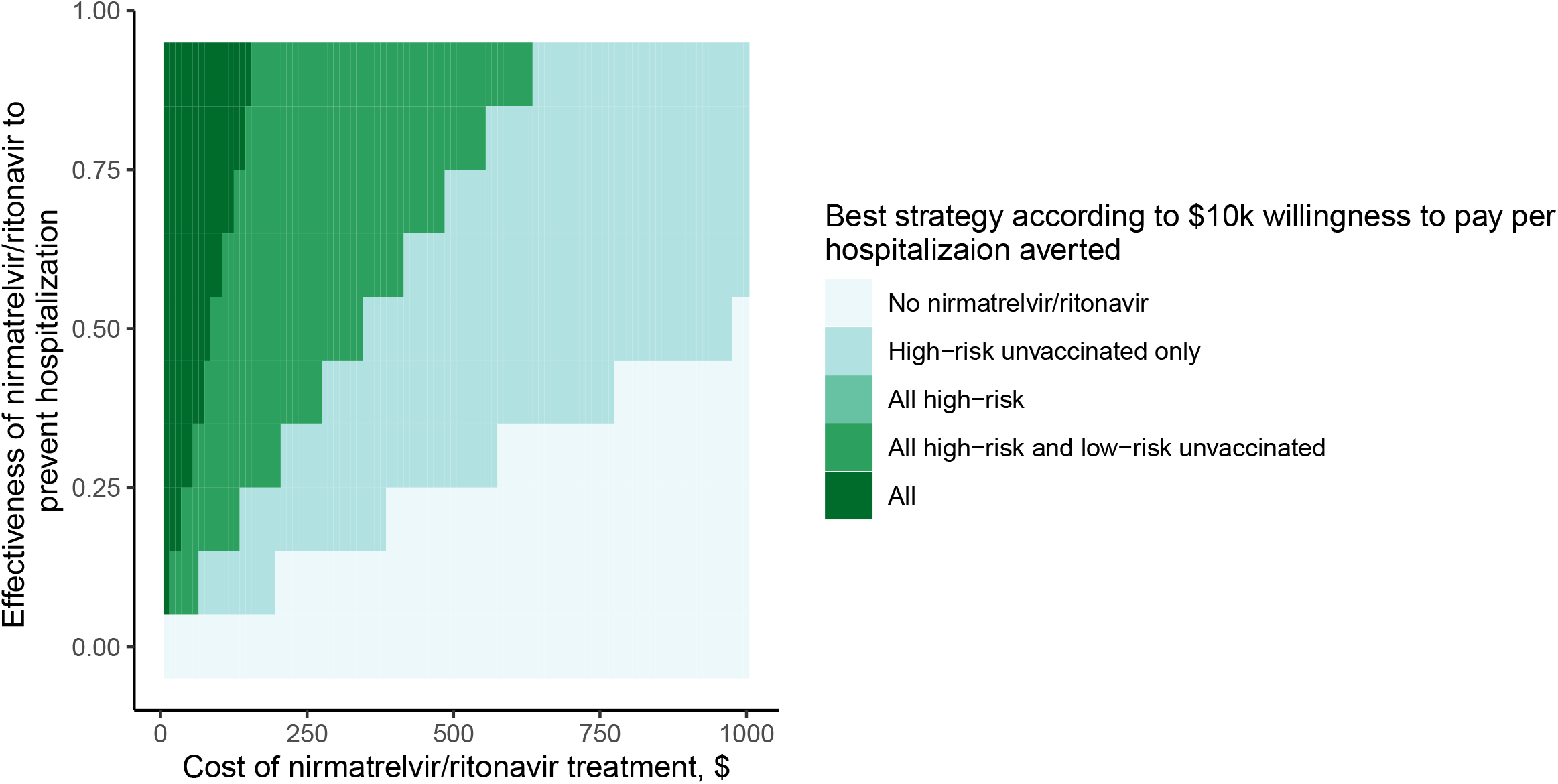
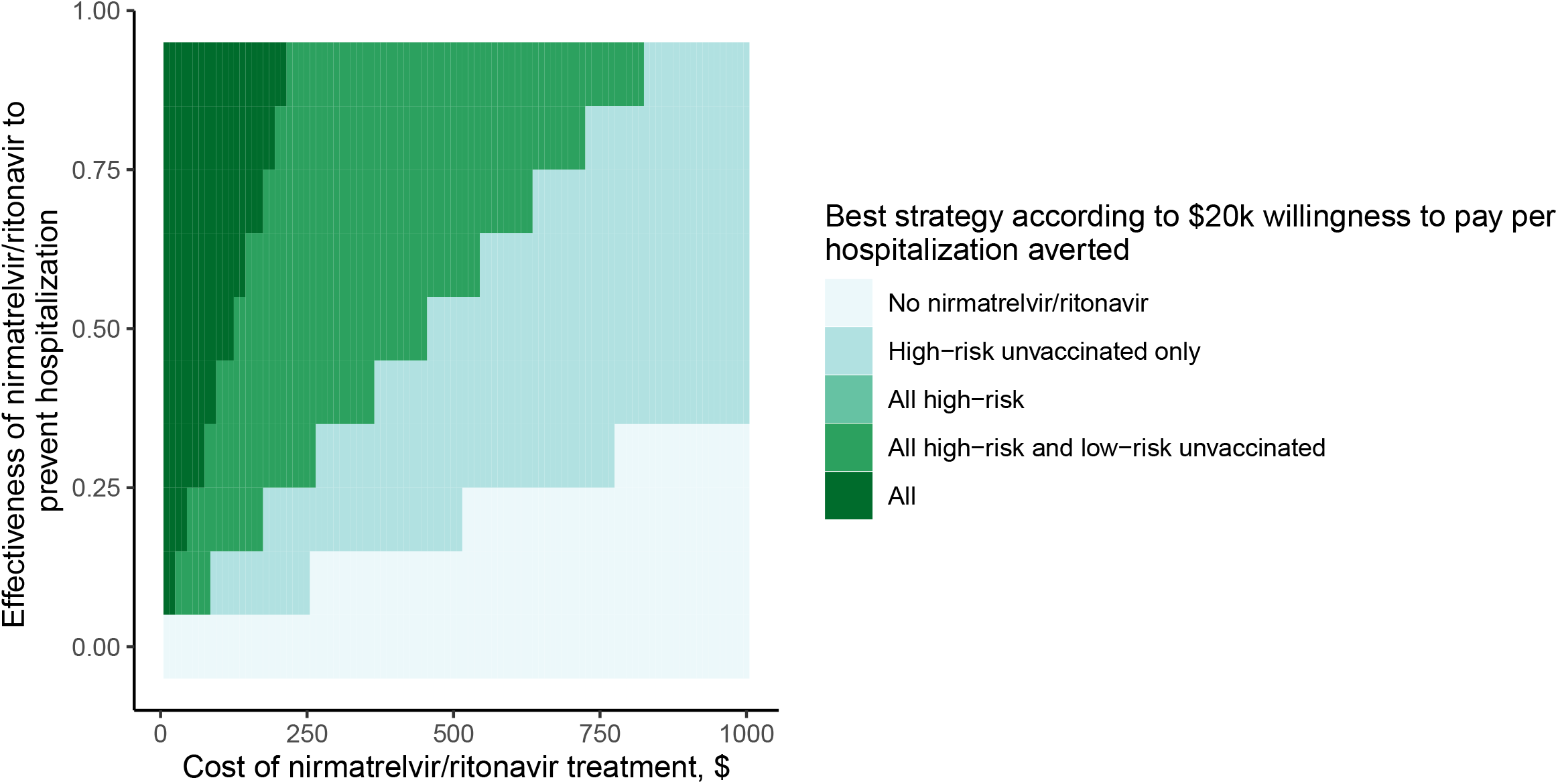
Two-way sensitivity analysis presenting most cost-effective treatment allocation strategy for hospitalizations averted, for given willingness-to-pay threshold, treatment effectiveness estimate, and cost of drug estimate. Cost of drug is on the horizontal axis, and treatment effectiveness is on the vertical axis.

**Figure 3.**
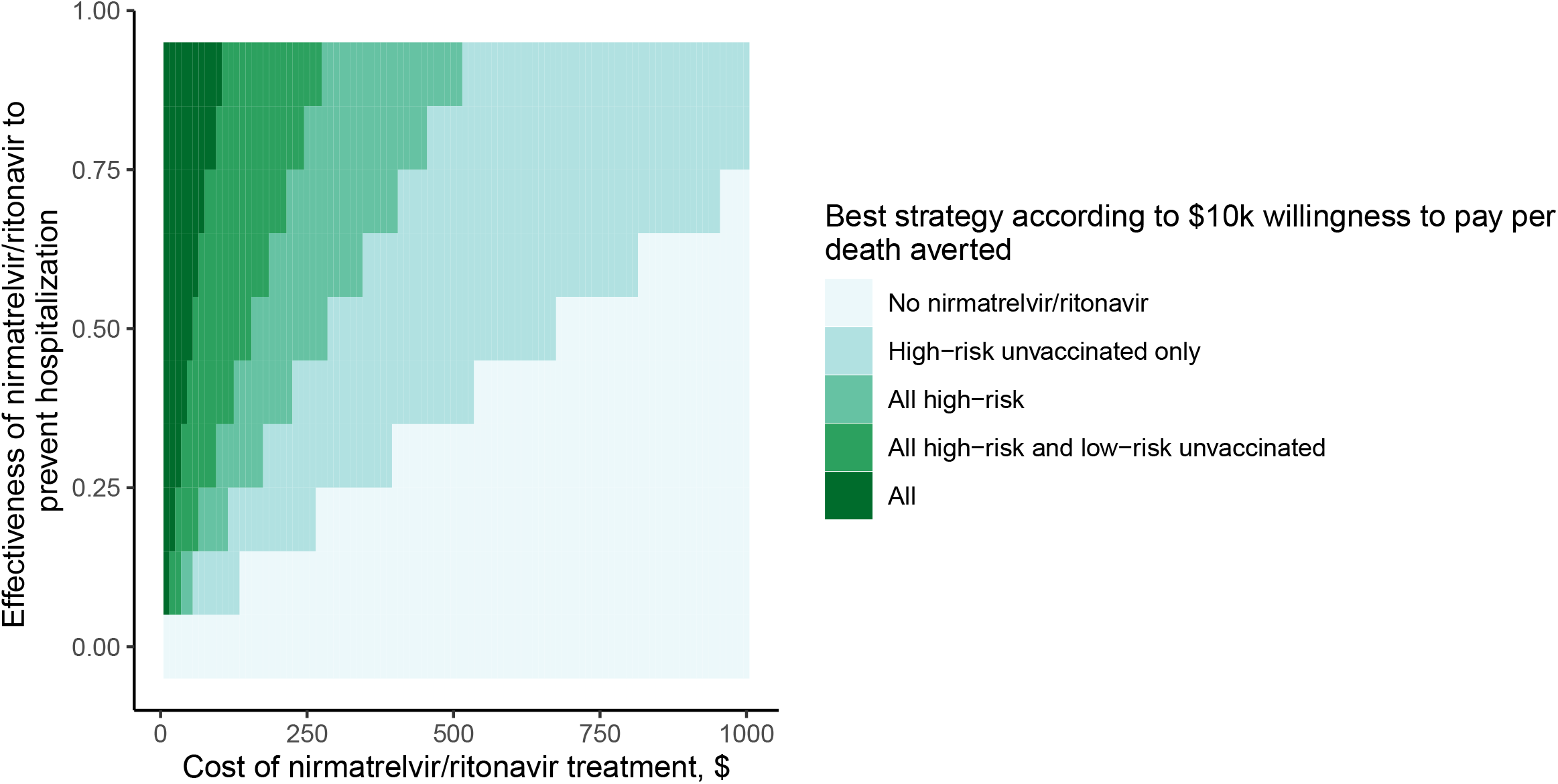
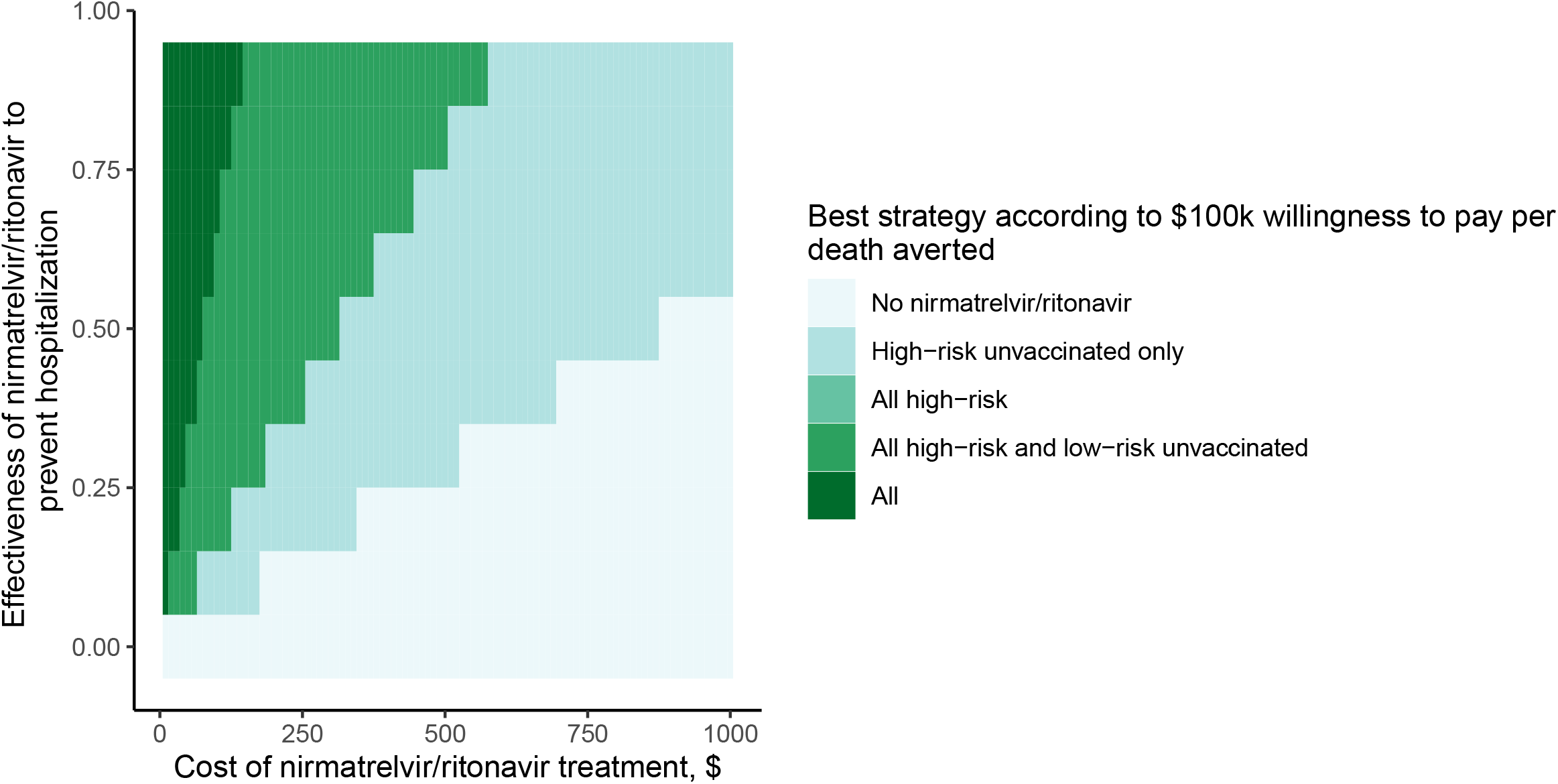
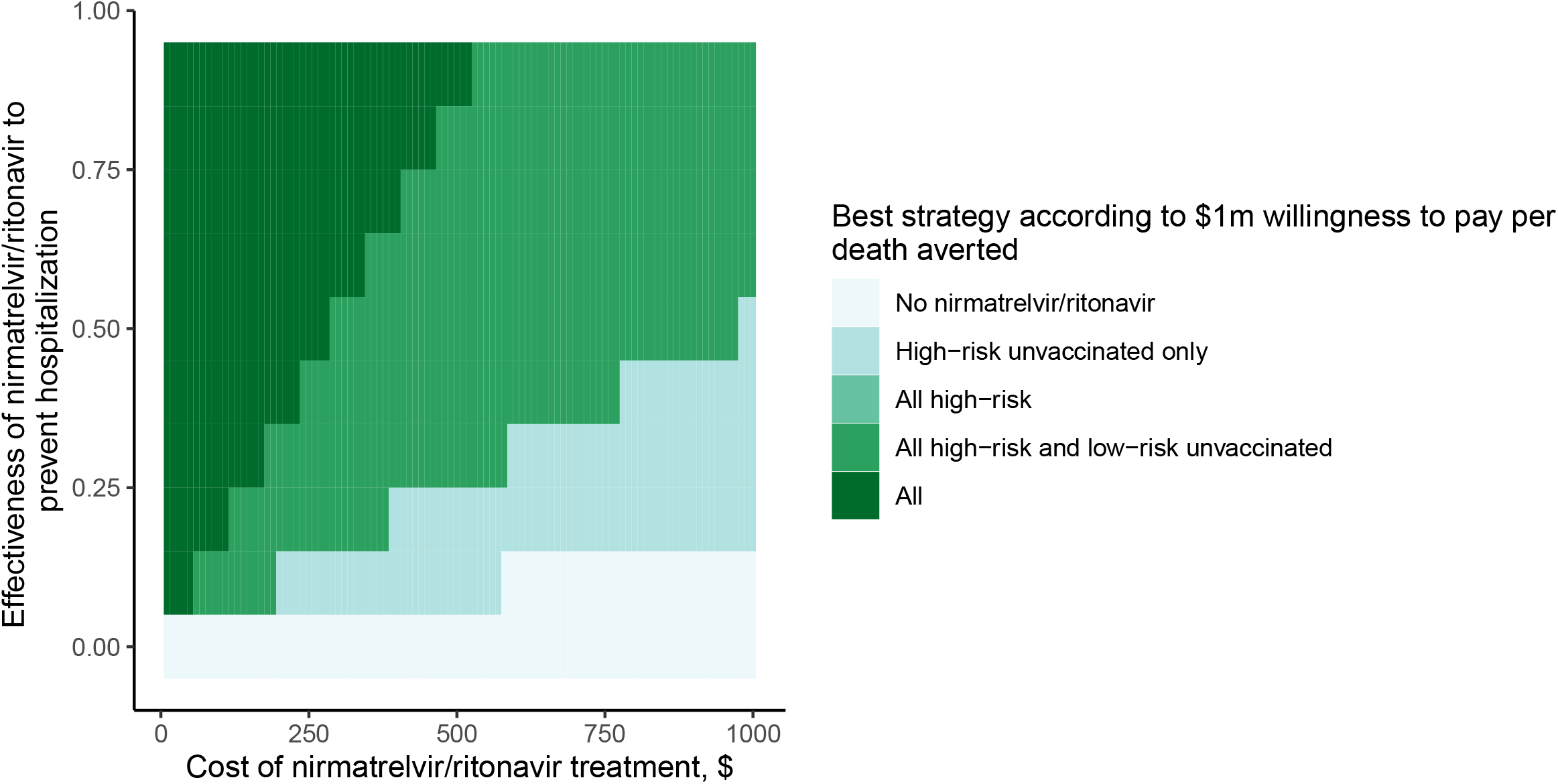
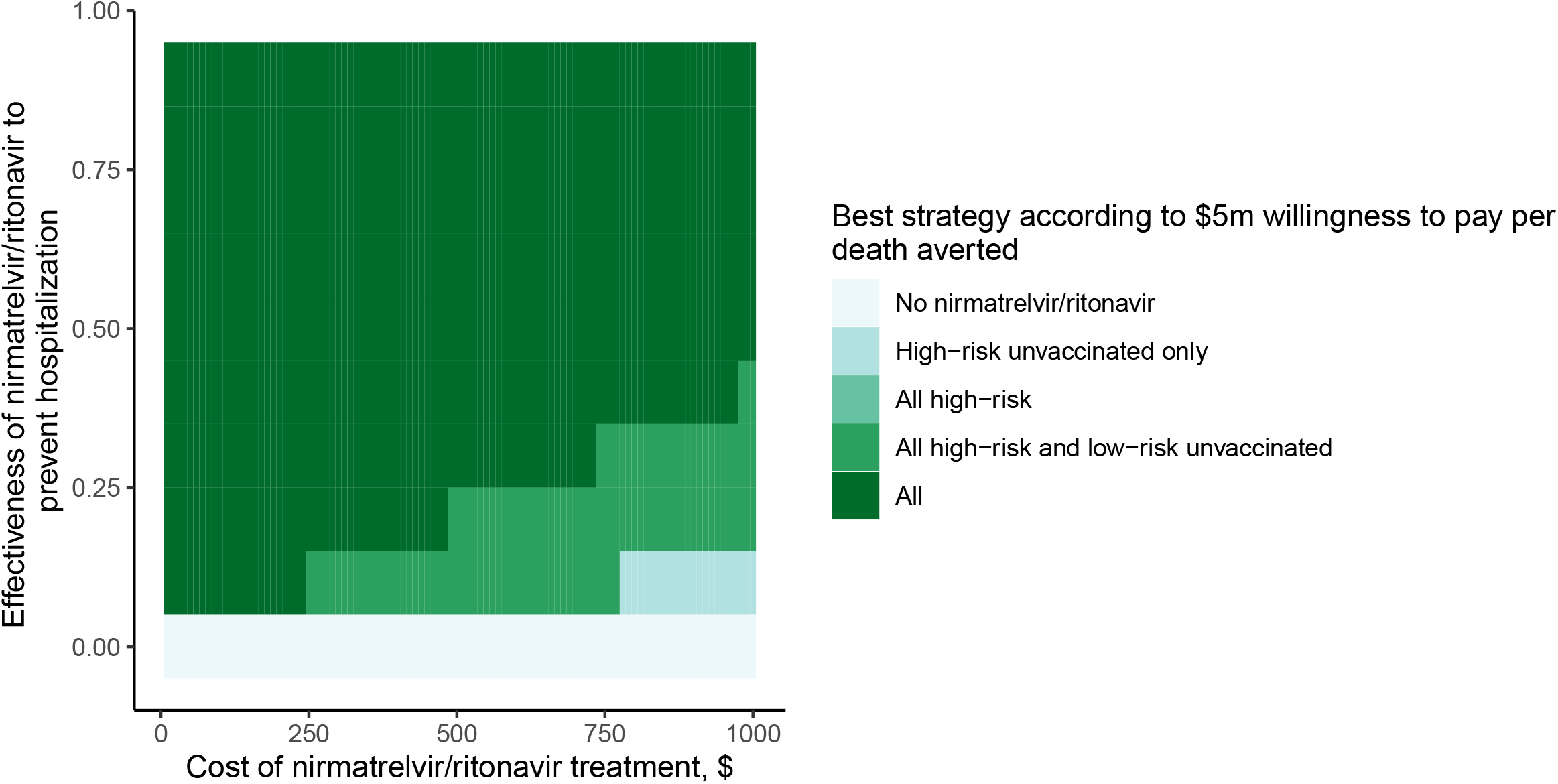
Two-way sensitivity analysis presenting most cost-effective treatment allocation strategy for deaths averted, for given willingness-to-pay threshold, treatment effectiveness estimate, and cost of drug estimate. Cost of drug is on the horizontal axis, and treatment effectiveness is on the vertical axis.

## Discussion

Several conclusions emerge from both the paradigmatic examples presented in the sections above and the countless alternative data scenarios that can be examined using the publicly available companion tool. First, unvaccinated persons at high-risk of severe COVID-19 should always have priority access to treatment, a strategy that was cost-saving across virtually every scenario we examined. Second, one size does not fit all in the subsequent assignment of priority: no single, “optimal” allocation plan captures the breadth of epidemiologic and drug performance circumstances. The curves presented in the Results section are malleable and dependent on the interplay of drug effectiveness, the comparative costs of both medications and hospitalization, the variant-specific risk of severe disease, and the societal willingness to pay to avert hospitalizations and deaths. Decision makers can and should tailor their allocation strategies to their particular settings. The web-based companion app is available to support such an exercise.

It is difficult to determine the societal willingness-to-pay to avert hospitalization and death. One frequently used measure is the Value of a Statistical Life (VSL). In the US, VSL estimates center around $10 million dollars per life (with low and high values between $5 million and $16 million) [23]. Since decision-makers may not feel comfortable with such a high willingness-to-pay threshold, we have offered a range of values here and a <u>webtool</u> that supports further exploration of alternative scenarios.

Our analysis aims to establish a priori standards for priority setting in anticipation of future developments in COVID-19 treatment. We use nirmatrelvir/ritonavir as an illustrative example but, in the spirit of exploration, we allowed its effectiveness to range widely, from the clinical trial estimate of 89% to a low of 21%. This reflects evidence that individual risk-level, vaccination status, and time of treatment initiation will likely influence treatment effectiveness [24]. It also acknowledges real-world studies suggesting reduced effectiveness with the newer Omicron variant [4, 7–10] and potentially reduced effectiveness in low-risk individuals [10]. In addition, there is evidence of people re-testing positive after their course of nirmatrelvir/ritonavir was completed, accompanied by a resurgence of symptoms, something that had not been seen in trials [14]. Finally, barriers to access (e.g., mandatory PCR testing and a physician visit to receive a prescription) will delay or impede treatment—particularly among poor and socially disadvantaged patients of color who are more likely to be at higher-risk for severe disease—further reducing effectiveness and exacerbating the inequities of COVID-19 care [25]. For all these reasons, our cost-effectiveness estimates should be interpreted as a best-case scenario for nirmatrelvir/ritonavir.

COVID-19 remains a global pandemic and treatments are needed worldwide. In the US, where a COVID-19 hospitalization can cost $11,000 to over $98,000, depending on its complexity [18, 19, 26], it is comparatively easy to establish the cost-effectiveness of a $500 course of treatment with nirmatrelvir/ritonavir. In low-income countries, hospitalization costs of $35 for a severe case and $310 for a critical case [27] make it much more difficult to justify the $500 cost of nirmatrelvir/ritonavir. Drug pricing for nirmatrelvir/ritonavir must be both context-specific and structured not to impede access to treatment. In a welcome development, Pfizer has pledged to offer nirmatrelvir/ritonavir (along with 22 additional medications) at non-profit prices for low-income countries [28]. Through work with the Clinton Health Access Initiative, a generic formulation of nirmatrelvir/ritonavir will be offered to low-income countries for $25 a course [29], though this leaves out middle income countries with high burden of COVID-19.

Our analysis has limitations. It does not consider drug supply or budgetary constraints, both of which might play a significant role in determining treatment allocation for decision-makers. We assume perfect adherence of those treated with the drug, which is likely unrealistic in a real-world scenario. However, without data to inform whether certain groups (vaccinated vs unvaccinated, high risk vs low risk) had different levels of adherence, we chose to keep this assumption with the understanding that it might bias our results towards groups with lower real-world adherence. In addition, we do not consider those who are contraindicated from taking nirmatrelvir/ritonavir due to other medications in this analysis. We ignore potential side-effects of treatment and drug-drug interactions, both of which may arise in clinical practice. We also ignore the possibility that patients may be inelgibile for nirmatrelvir/ritonavir due to other medications, which might preclude a large percentage of the highest-risk population and prevent a large percentage of the benefit seen in this analysis. Our model does not consider disease transmission, nor does it account for any costs other than hospitalization and nirmatrelvir/ritonavir treatment. Specifically, we do not consider the costs of either PCR testing to confirm a diagnosis of COVID (required for treatment in the U.S.) or a physician’s office visit to be evaluated and receive a prescription.

In conclusion, our adaptable quantitative framework demonstrates that for almost every scenario evaluating appropriate treatment allocation, prescribing nirmatrelvir/ritonavir to unvaccinated patients at high-risk of severe COVID-19 was cost-saving, meaning this group should almost always be treated if treatment is available. In other strategies, the most cost-effective allocation of nirmatrelvir/ritonavir can be determined via a formal weighing of a variety of factors, including treatment effectiveness, both overall and within allocation groups, and the drug’s cost. Such a framework can help decision-makers choose the most appropriate allocation strategy given changing information and disease dynamics.

## Data Availability

All data is publicly available at

https://github.com/ASavinkina/COVIDantiviral

https://savinkina.shinyapps.io/Paxlovid/

## References

1. Hannah Ritchie EM, Lucas Rodés-Guirao, Cameron Appel, Charlie Giattino, Esteban Ortiz-Ospina, Joe Hasell, Bobbie Macdonald, Diana Beltekian and Max Roser. Coronavirus Pandemic (COVID-19). Published online at OurWorldInData.org, 2020.

2. Jared Ortaliza KA, Cynthia Cox. COVID-19 leading cause of death ranking. Peterson-KFF Health System Tracker, 2022.

3. Eric C. Schneider AS, Pratha Sah, Thomas Vilches, Abhishek Pandey, Seyed M. Moghadas, Alison Galvani. Impact of U.S. COVID-19 Vaccination Efforts: An Update on Averted Deaths, Hospitalizations, and Health Care Costs Through March 2022. To the Point (blog). Commonwealth Fund, 2022.

4. Hammond J, Leister-Tebbe H, Gardner A, et al. Oral Nirmatrelvir for High-Risk, Nonhospitalized Adults with Covid-19. Commonwealth FundN Engl J Med 2022; 386(15): 1397–408.

5. Carl Zimmer KJW, Jonathan Corum, Matthew Kristoffersen. Coronavirus Drug and Treatment Tracker. Commonwealth FundThe New York Times. 2022 5/12/22.

6. PAXLOVID Fact Sheet for Healthcare Providers. Pfizer Inc, 2021.

7. Mefsin Y, Chen D, Bond HS, et al. Epidemiology of infections with SARS-CoV-2 Omicron BA.2 variant in Hong Kong, January-March 2022. medRxiv 2022: 2022.04.07.22273595.

8. Yip CFaL, Grace C.Y. and Man Lai, Mandy Sze and Wong, Vincent Wai-Sun and Tse, Yee-Kit and Ma Bosco Hon-Ming and Hui, Elsie and Leung Maria KW and Chan Henry Lik-Yuen and Hui David S. C. and Hui, David Shu-Cheong. Impact of the Use of Oral Antiviral Agents on the Risk of Hospitalisation in Community COVID-19 Patients. PrePrint Available at SSRN 2022.

9. Dryden-Peterson S, Kim A, Kim AY, et al. Nirmatrelvir plus ritonavir for early COVID-19 and hospitalization in a large US health system. medRxiv 2022: 2022.06.14.22276393.

10. Ronen Arbel YWS, Moshe Hoshen et al. Oral Nirmatrelvir and Severe Covid-19 Outcomes During the Omicron Surge. PREPRINT (Version 1) available at Research Square 2022.

11. Jarvis L. The U.S. Is Doing Too Little to Monitor Paxlovid Use. Bloomberg, 2022.

12. Fenyves P. Does Paxlovid help people who have been vaccinated against Covid-19? Show us the data! STAT, 2022.

13. Ryan B. Pfizer antiviral pills may be risky with other medications. NBC News, 2021.

14. Rubin R. From Positive to Negative to Positive Again-The Mystery of Why COVID-19 Rebounds in Some Patients Who Take Paxlovid. Jama 2022.

15. Beasley D. U.S. doctors reconsider Pfizer’s Paxlovid for lower-risk COVID patients. Reuters, 2022.

16. Sax PE. Should We Prescribe Nirmatrelvir/r (Paxlovid) to Low-Risk COVID-19 Patients? HIV and ID Observations Vol. 2022. NEJM Journal Watch, 2022.

17. Mishra M. U.S. to buy 10 mln courses of Pfizer’s COVID-19 pill for $5.3 bln. Reuters, 2021.

18. Tsai Y, Vogt TM, Zhou F. Patient Characteristics and Costs Associated With COVID-19-Related Medical Care Among Medicare Fee-for-Service Beneficiaries. Ann Intern Med 2021; 174(8): 1101–9.

19. Ohsfeldt RL, Choong CK, Mc Collam PL, Abedtash H, Kelton KA, Burge R. Inpatient Hospital Costs for COVID-19 Patients in the United States. Adv Ther 2021; 38(11): 5557–95.

20. Sanders GD, Neumann PJ, Basu A, et al. Recommendations for Conduct, Methodological Practices, and Reporting of Cost-effectiveness Analyses: Second Panel on Cost-Effectiveness in Health and Medicine. Jama 2016; 316(10): 1093–103.

21. Stinnett AA, Mullahy J. Net health benefits: a new framework for the analysis of uncertainty in cost-effectiveness analysis. Med Decis Making 1998; 18(2 Suppl): S68–80.

22. Landefeld JS, Seskin EP. The economic value of life: linking theory to practice. Am J Public Health 1982; 72(6): 555–66.

23. Office of the Assistant Secretary for Planning and Evaluation USDoHaHS. Guidelines for regulatory impact analysis. 2016. 2016.

24. Wong CKH, Au ICH, Lau KTK, Lau EHY, Cowling BJ, Leung GM. Real-world effectiveness of molnupiravir and nirmatrelvir/ritonavir among COVID-19 inpatients during Hong Kong’s Omicron BA.2 wave: an observational study. medRxiv 2022: 2022.05.19.22275291.

25. Vasan A, Foote M, Long T. Ensuring Widespread and Equitable Access to Treatments for COVID-19. JAMA 2022.

26. FairHealth. National Average Charge for a Complex Hospital Stay for COVID-19 Is $317,810, FAIR Health Finds. 2021.

27. Torres-Rueda S, Sweeney S, Bozzani F, et al. Stark choices: exploring health sector costs of policy responses to COVID-19 in low-income and middle-income countries. BMJ Glob Health 2021; 6(12).

28. Mullin R. Pfizer, ViiV launch drug programs for low-income countries. Chemical and Engineering News, 2022.

29. CHAI. Press release: CHAI announces agreements with leading generic manufacturers to make affordable COVID-19 treatment available in low- and middle-income countries. Clinton Health Access Initiative, 2022.

30. Bureau USC. National Demographic Analysis Tables: 2020, 2022.

31. Koma WN, Tracy; Claxton, Gary; Rae, Matthew; Kates, Jennifer; Michaud, Josh. How Many Adults Are at Risk of Serious Illness If Infected with Coronavirus? Updated Data: Henry J. Kaiser Family Foundation, 2020 4/23/2020.

32. Adams ML, Katz DL, Grandpre J. Population-Based Estimates of Chronic Conditions Affecting Risk for Complications from Coronavirus Disease, United States. Emerg Infect Dis 2020; 26(8): 1831–3.

33. Ajufo E, Rao S, Navar AM, Pandey A, Ayers CR, Khera A. U.S. population at increased risk of severe illness from COVID-19. Am J Prev Cardiol 2021; 6: 100156.

34. Pfizer. EPIC-HR: Study of Oral PF-07321332/Ritonavir Compared With Placebo in Nonhospitalized High Risk Adults With COVID-19. 2022.

35. Jehi L, Ji X, Milinovich A, et al. Development and validation of a model for individualized prediction of hospitalization risk in 4,536 patients with COVID-19. PLoS One 2020; 15(8): e0237419.

36. CDC. Risk for COVID-19 Infection, Hospitalization, and Death by Age Group, 2022 3/28/2022.

37. Taylor CA, Whitaker M, Anglin O, et al. COVID-19-Associated Hospitalizations Among Adults During SARS-CoV-2 Delta and Omicron Variant Predominance, by Race/Ethnicity and Vaccination Status - COVID-NET, 14 States, July 2021-January 2022. MMWR Morb Mortal Wkly Rep 2022; 71(12): 466–73.

38. CDC. COVID Data Tracker. Atlanta, GA: US: Department of Health and Human Services, CDC, 2022 4/21/2022.

