## Supplemental Tables for "Determining population-level allocation strategies for COVID-19 treatments in the United States using a quantitative framework, a case study using nirmatrelvir/ritonavir"

**Supplemental Table 1. Initial hospitalization results.** Cost effectiveness table assessing average per-infection

costs of providing nirmatrelvir/ritonavir, average per-infection risk of hospitalization, and incremental cost-effectiveness ratio for cost-per-hospitalization averted. Panels show differing effectiveness measures against hospitalization from literature: A. 89%, Hammond et al, B. 21%, Yip et al. C. 67%, Arbel et al D. 45%, Dryden-Peterson, E. 33%, Mefsin et al.^5^

**A.**

|  | Probability Hospitalizations, per infection | Average Cost, per infection | ICER, per hospitalization averted |
| --- | --- | --- | --- |
| Strategy 0: No nirmatrelvir/ritonavir | 0.0268 | $536.00 |  |
| Strategy 1 - nirmatrelvir/ritonavir for unvaccinated high-risk | 0.0152 | $413.00 | Cost saving |
| Strategy 2 - nirmatrelvir/ritonavir for all high-risk (regardless of vaccination) | 0.0058 | $423.00 | $700 |
| Strategy 3 - nirmatrelvir/ritonavir all high risk and unvaccinated low-risk | 0.0042 | $465.00 | $26,700 |
| Strategy 4 - nirmatrelvir/ritonavir for all | 0.0030 | $577.00 | $88,600 |

| B. |  |  |  |
| --- | --- | --- | --- |
|  | **Probability Hospitalizations, per infection** | **Average Cost, per infection** | **ICER, per hospitalization averted** |
| Strategy 0: No nirmatrelvir/ritonavir | 0.0268 | $536.00 |  |
| Strategy 1 - nirmatrelvir/ritonavir for unvaccinated high-risk | 0.0241 | $590.00 | $19,530 |
| Strategy 2 - nirmatrelvir/ritonavir for all high-risk (regardless of vaccination) | 0.0219 | $745.00 | $69,300 |
| Strategy 3 - nirmatrelvir/ritonavir all high risk and unvaccinated low-risk | 0.0215 | $810.00 | $179,800 |
| Strategy 4 - nirmatrelvir/ritonavir for all | 0.0212 | $942.00 | $441,900 |

| C. |  |  |  |
| --- | --- | --- | --- |
|  | **Probability Hospitalizations, per infection** | **Average Cost, per infection** | **ICER, per hospitalization averted** |
| Strategy 0: No nirmatrelvir/ritonavir | 0.0268 | $536.00 |  |
| Strategy 1 - nirmatrelvir/ritonavir for unvaccinated high-risk | 0.0181 | $467.00 | Cost-Saving |
| Strategy 2 - nirmatrelvir/ritonavir for all high-risk (regardless of vaccination) | 0.0110 | $521.00 | $7,622 |
| Strategy 3 - nirmatrelvir/ritonavir all high risk and unvaccinated low-risk | 0.0098 | $570.00 | $42,300 |
| Strategy 4 - nirmatrelvir/ritonavir for all | 0.0089 | $689.00 | $124,400 |

| D. |  |  |  |
| --- | --- | --- | --- |
|  | **Probability Hospitalizations, per infection** | **Average Cost, per infection** | **ICER, per hospitalization averted** |
| Strategy 0: No nirmatrelvir/ritonavir | 0.0268 | $536.00 |  |
| Strategy 1 - nirmatrelvir/ritonavir for unvaccinated high-risk | 0.0210 | $526.00 | Cost-Saving |
| Strategy 2 - nirmatrelvir/ritonavir for all high-risk (regardless of vaccination) | 0.0162 | $628.00 | $21,400 |
| Strategy 3 - nirmatrelvir/ritonavir all high risk and unvaccinated low-risk | 0.0154 | $685.00 | $72,900 |
| Strategy 4 - nirmatrelvir/ritonavir for all | 0.0148 | $810.00 | $195,300 |

**E.**

|  | Probability Hospitalization, per infection | Average Cost, per infection | ICER, per hospitalization averted |
| --- | --- | --- | --- |
| Strategy 0: No nirmatrelvir/ritonavir | 0.0268 | $536.00 |  |
| Strategy 1 - nirmatrelvir/ritonavir for unvaccinated high-risk | 0.0225 | $558.00 | $4,963 |
| Strategy 2 - nirmatrelvir/ritonavir for all high-risk (regardless of vaccination) | 0.0190 | $687.00 | $36,600 |
| Strategy 3 - nirmatrelvir/ritonavir all high risk and unvaccinated low-risk | 0.0184 | $748.00 | $106,900 |
| Strategy 4 - nirmatrelvir/ritonavir for all | 0.0180 | $876.00 | $273,800 |

**Supplemental Table 2. Initial deaths results.** Cost effectiveness table assessing average per-infection costs of providing nirmatrelvir/ritonavir, average per-infection risk of death, and incremental cost-effectiveness ratio for cost-per-death averted. Panels show differing effectiveness measures against hospitalization from literature: A. 89%, Hammond et al,^1^ B. 21%, Yip et al.^2^ C. 67%, Arbel et al^3^ D. 45%, Dryden-Peterson^4^, E. 33%, Mefsin et al.^5^

**A.**

|  | Probability Death, per infection | Average Cost, per infection | ICER, per death averted |
| --- | --- | --- | --- |
| Strategy 0: No nirmatrelvir/ritonavir | 0.00187 | $536.00 |  |
| Strategy 1 - nirmatrelvir/ritonavir for unvaccinated high-risk | 0.00107 | $413.00 | Cost saving |
| Strategy 2 - nirmatrelvir/ritonavir for all high-risk (regardless of vaccination) | 0.00040 | $423.00 | $9,500 |
| Strategy 3 - nirmatrelvir/ritonavir all high risk and unvaccinated low-risk | 0.00030 | $465.00 | $381,900 |
| Strategy 4 - nirmatrelvir/ritonavir for all | 0.00021 | $577.00 | $1,265,600 |
| **B.** |  |  |  |
|  | **Probability Death, per infection** | **Average Cost, per infection** | **ICER, per death averted** |
| Strategy 0: No nirmatrelvir/ritonavir | 0.00187 | $536.00 |  |
| Strategy 1 - nirmatrelvir/ritonavir for unvaccinated high-risk | 0.00169 | $590.00 | $279,000 |
| Strategy 2 - nirmatrelvir/ritonavir for all high-risk (regardless of vaccination) | 0.00153 | $745.00 | $989,800 |
| Strategy 3 - nirmatrelvir/ritonavir all high risk and unvaccinated low-risk | 0.00150 | $810.00 | $2,568,200 |
| Strategy 4 - nirmatrelvir/ritonavir for all | 0.00148 | $942.00 | $6,313,500 |

| C. |  |  |  |
| --- | --- | --- | --- |
|  | **Probability Death, per infection** | **Average Cost, per infection** | **ICER, per death averted** |
| Strategy 0: No nirmatrelvir/ritonavir | 0.00187 | $536.00 |  |
| Strategy 1 - nirmatrelvir/ritonavir for unvaccinated high-risk | 0.00127 | $467.00 | Cost-Saving |
| Strategy 2 - nirmatrelvir/ritonavir for all high-risk (regardless of vaccination) | 0.00077 | $521.00 | $108,900 |
| Strategy 3 - nirmatrelvir/ritonavir all high risk and unvaccinated low-risk | 0.00069 | $570.00 | $603,600 |
| Strategy 4 - nirmatrelvir/ritonavir for all | 0.00062 | $689.00 | $1,777,500 |
| D. |  |  |  |
|  | **Probability Death, per infection** | **Average Cost, per infection** | **ICER, per death averted** |
| Strategy 0: No nirmatrelvir/ritonavir | 0.00187 | $536.00 |  |
| Strategy 1 - nirmatrelvir/ritonavir for unvaccinated high-risk | 0.00147 | $526.00 | Cost-Saving |
| Strategy 2 - nirmatrelvir/ritonavir for all high-risk (regardless of vaccination) | 0.00113 | $628.00 | $305,500 |
| Strategy 3 - nirmatrelvir/ritonavir all high risk and unvaccinated low-risk | 0.00108 | $685.00 | $1,042,100 |
| Strategy 4 - nirmatrelvir/ritonavir for all | 0.00103 | $810.00 | $2,789,900 |

**E.**

|  | Probability Death, per infection | Average Cost, per infection | ICER, per death averted |
| --- | --- | --- | --- |
| Strategy 0: No nirmatrelvir/ritonavir | 0.00187 | $536.00 |  |
| Strategy 1 - nirmatrelvir/ritonavir for unvaccinated high-risk | 0.00158 | $558.00 | $70,900 |
| Strategy 2 - nirmatrelvir/ritonavir for all high-risk (regardless of vaccination) | 0.00133 | $687.00 | $523,200 |
| Strategy 3 - nirmatrelvir/ritonavir all high risk and unvaccinated low-risk | 0.00129 | $748.00 | $1,527,600 |
| Strategy 4 - nirmatrelvir/ritonavir for all | 0.00126 | $876.00 | $3,911,000 |

**Supplemental Table 3. Sensitivity analysis with differential nirmatrelvir/ritonavir effectiveness by risk status for severe COVID-19.** By differing allocation scenarios, and presented for different effectiveness at preventing hospitalization estimates for nirmatrelvir/ritonavir from the literature. Effectiveness for high-risk people as listed, and for low-risk at 33% that of high-risk, based off Arbel at al. study which showed 67% effectiveness in high-risk people and 22% effectiveness in low-risk.

|  | Incremental cost-effectiveness ratios (ICER) per hospitalization averted, according to given nirmatrelvir/ritonavir effectiveness at preventing hospitalization measure | | | | |
| --- | --- | --- | --- | --- | --- |
|  | 89% effective,  Hammond et al.^6^ | 67% effective, Arbel et al.^12^ | 45% effective,  Dryden-Peterson et al.^11^ | 33% effective,  Mefsin et al.^9^ | 21% effective,  Yip et al.^10^ |
| Strategy 0: No nirmatrelvir/ritonavir |  |  |  |  |  |
| Strategy 1 - nirmatrelvir/ritonavir for unvaccinated high-risk | Cost saving | Cost-Saving | Cost-Saving | $5,000 | $19,500 |
| Strategy 2 - nirmatrelvir/ritonavir for all high-risk (regardless of vaccination) | $700 | $7,600 | $21,400 | $36,600 | $69,300 |
| Strategy 3 - nirmatrelvir/ritonavir all high risk and unvaccinated low-risk | $122,700 | $169,700 | $262,700 | $365,700 | $586,400 |
| Strategy 4 - nirmatrelvir/ritonavir for all | $310,100 | $418,700 | $633,500 | $871,300 | $1,380,900 |

**Supplemental Table 4. Sensitivity analysis with differential nirmatrelvir/ritonavir effectiveness by vaccination status.** By differing allocation scenarios, and presented for different effectiveness at preventing hospitalization estimates for nirmatrelvir/ritonavir from the literature. Effectiveness for unvaccinated people as listed, and for vaccinated at 50% that of unvaccinated.

|  | Incremental cost-effectiveness ratios (ICER) per hospitalization averted, according to given nirmatrelvir/ritonavir effectiveness at preventing hospitalization measure | | | | |
| --- | --- | --- | --- | --- | --- |
|  | 89% effective,  Hammond et al.^6^ | 67% effective, Arbel et al.^12^ | 45% effective,  Dryden-Peterson et al.^11^ | 33% effective,  Mefsin et al.^9^ | 21% effective,  Yip et al.^10^ |
| Strategy 0: No nirmatrelvir/ritonavir |  |  |  |  |  |
| Strategy 1 - nirmatrelvir/ritonavir for unvaccinated high-risk | Cost saving | Cost-Saving | Cost-Saving | $5,000 | $19,500 |
| Strategy 2 - nirmatrelvir/ritonavir for all high-risk (regardless of vaccination) | $1,300 | $15,200 | $42,800 | $73,300 | $138,600 |
| Strategy 3 - nirmatrelvir/ritonavir all high risk and unvaccinated low-risk | $26,700 | $42,300 | $72,900 | $106,900 | $179,800 |
| Strategy 4 - nirmatrelvir/ritonavir for all | $351,700 | $423,400 | $565,100 | $722,000 | $1,058,400 |

**Supplemental Table 5. Sensitivity analysis with differential cost per average COVID hospitalization.** By differing allocation scenarios, and presented for different effectiveness at preventing hospitalization estimates for nirmatrelvir/ritonavir from the literature. Average cost per US COVID-19 hospitalization changed from $20,000 to $10,000.

|  | Incremental cost-effectiveness ratios (ICER) per hospitalization averted, according to given nirmatrelvir/ritonavir effectiveness at preventing hospitalization measure | | | | |
| --- | --- | --- | --- | --- | --- |
|  | 89% effective,  Hammond et al.^6^ | 67% effective, Arbel et al.^12^ | 45% effective,  Dryden-Peterson et al.^11^ | 33% effective,  Mefsin et al.^9^ | 21% effective,  Yip et al.^10^ |
| Strategy 0: No nirmatrelvir/ritonavir |  |  |  |  |  |
| Strategy 1 - nirmatrelvir/ritonavir for unvaccinated high-risk | Cost saving | $2,000 | $8,200 | $15,000 | $29,500 |
| Strategy 2 - nirmatrelvir/ritonavir for all high-risk (regardless of vaccination) | $10,700 | $17,600 | $31,400 | $46,600 | $79,200 |
| Strategy 3 - nirmatrelvir/ritonavir all high risk and unvaccinated low-risk | $36,700 | $52,300 | $82,900 | $116,900 | $189,800 |
| Strategy 4 - nirmatrelvir/ritonavir for all | $98,600 | $134,400 | $205,300 | $283,800 | $451,900 |

**Supplemental Table 6. Sensitivity analysis with increased risk of hospitalization from COVID-19.** By differing allocation scenarios and presented for different effectiveness at preventing hospitalization estimates for nirmatrelvir/ritonavir from the literature. The risk of hospitalization is doubled to account for a hypothetically more severe variant.

|  | Incremental cost-effectiveness ratios (ICER) per hospitalization averted, according to given nirmatrelvir/ritonavir effectiveness at preventing hospitalization measure | | | | |
| --- | --- | --- | --- | --- | --- |
|  | 89% effective,  Hammond et al.^6^ | 67% effective, Arbel et al.^12^ | 45% effective,  Dryden-Peterson et al.^11^ | 33% effective,  Mefsin et al.^9^ | 21% effective,  Yip et al.^10^ |
| Strategy 0: No nirmatrelvir/ritonavir |  |  |  |  |  |
| Strategy 1 - nirmatrelvir/ritonavir for unvaccinated high-risk | Cost saving | Cost saving | Cost saving | $2,200 | $9,500 |
| Strategy 2 - nirmatrelvir/ritonavir for all high-risk (regardless of vaccination) | Cost saving | $3,000 | $9,600 | $17,000 | $32,700 |
| Strategy 3 - nirmatrelvir/ritonavir all high risk and unvaccinated low-risk | $3,100 | $20,900 | $36,200 | $53,200 | $89,600 |
| Strategy 4 - nirmatrelvir/ritonavir for all | $33,600 | $61,400 | $96,600 | $135,600 | $219,000 |

**References**

1.	Hammond J, Leister-Tebbe H, Gardner A, et al. Oral Nirmatrelvir for High-Risk, Nonhospitalized Adults with Covid-19. *N Engl J Med* 2022; **386**(15): 1397-408.

2.	Yip CFaL, Grace C.Y. and Man Lai, Mandy Sze and Wong, Vincent Wai-Sun and Tse, Yee-Kit and Ma, Bosco Hon-Ming and Hui, Elsie and Leung, Maria KW and Chan, Henry Lik-Yuen and Hui, David S. C. and Hui, David Shu-Cheong. Impact of the Use of Oral Antiviral Agents on the Risk of Hospitalisation in Community COVID-19 Patients. *PrePrint Available at SSRN* 2022.

3.	Ronen Arbel YWS, Moshe Hoshen et al. . Oral Nirmatrelvir and Severe Covid-19 Outcomes During the Omicron Surge. *PREPRINT (Version 1) available at Research Square* 2022.

4.	Dryden-Peterson S, Kim A, Kim AY, et al. Nirmatrelvir plus ritonavir for early COVID-19 and hospitalization in a large US health system. *medRxiv* 2022: 2022.06.14.22276393.

5.	Mefsin Y, Chen D, Bond HS, et al. Epidemiology of infections with SARS-CoV-2 Omicron BA.2 variant in Hong Kong, January-March 2022. *medRxiv* 2022: 2022.04.07.22273595.
